# Serological survey of SARS-CoV-2 incidence conducted at a rural West Virginia hospital

**DOI:** 10.1101/2021.08.16.21262128

**Authors:** Alexander M. Horspool, Brynnan P. Russ, M. Allison Wolf, Jason Kang, Catherine B. Blackwood, Jesse M. Hall, Ting Y. Wong, Megan A. DeJong, Graham Bitzer, Justin R. Bevere, Robert Eggleston, Anita Stewart, Lisa Costello, Shelley Welch, Theodore Kieffer, Sally Hodder, F. Heath Damron

## Abstract

The SARS-CoV-2 pandemic has affected all types of global communities. Differences in urban and rural environments have led to varying levels of transmission within these subsets of the population. To fully understand the prevalence and impact of SARS-CoV-2 it is critical to survey both types of community. This study establishes the prevalence of SARS-CoV-2 in a rural community: Montgomery, West Virginia. Approximately 10% of participants exhibited serological or PCR-based results indicating exposure to SARS-CoV-2 within 6 months of the sampling date. Quantitative analysis of IgG levels against SARS-CoV-2 receptor binding domain (RBD) was used to stratify individuals based on antibody response to SARS-CoV-2. A significant negative correlation between date of exposure and degree of anti-SARS-CoV-2 RBD IgG (R^2^ = 0.9006) was discovered in addition to a correlation between neutralizing anti-SARS-CoV-2 antibodies (R^2^ = 0.8880) and days post exposure. Participants were confirmed to have normal immunogenic profiles by determining serum reactivity *B. pertussis* antigens commonly used in standardized vaccines. No significant associations were determined between anti-SARS-CoV-2 RBD IgG and age or biological sex. Reporting of viral-like illness symptoms was similar in SARS-CoV-2 exposed participants greater than 30 years old (100% reporting symptoms 30-60 years old, 75% reporting symptoms >60 years old) in contrast to participants under 30 years old (25% reporting symptoms). Overall, this axnalysis of a rural population provides important information about the SARS-CoV-2 pandemic in small rural communities. The study also underscores the fact that prior infection with SARS-CoV-2 results in antibody responses that wane over time which highlights the need for vaccine mediated protection in the absence of lasting protection.

## INTRODUCTION

SARS-CoV-2 and the COVID-19 pandemic has dramatically impacted public health across the globe. In the early phase of the pandemic, SARS-CoV-2 rapidly infected urban centers in California, New York, and other major U.S. cities while infecting rural communities at reduced rates (Anand et al., 2020; Bajema et al., 2020; Duca et al., 2020; Menachemi et al., 2020). High infection rates in urban areas continued until July of 2020 at which point relative incidence began to decline. In September of 2020, SARS-CoV-2 prevalence surged, this time including rural communities (Bajema et al., 2020; Duca et al., 2020). West Virginia was one of the last remaining states to report an active SARS-CoV-2 infection. This was likely due to a combination of early preventative measures and its highly rural population in contrast to some major urban locations. However, several states, including West Virginia, were dramatically impacted by the second surge after September 2020 and may have unique SARS-CoV-2 infection dynamics (Anand et al., 2020). As such, it is important to understand the impact of SARS-CoV-2 in these populations.

Rural communities comprise approximately 95% of the land mass of the United States but only 19.3% of the total population (Health Resources & Services Administration, 2021). Population densities in these areas are vastly different than major urban centers, often having an average of 45 times fewer people per square mile than major urban centers (Cohen, 2015). The impact on these communities during infectious disease outbreaks has been discussed for years. Specifically, rural communities have considerably different epidemic transmission concerns. Geographically disperse populations result in greater journeys to medical facilities or testing sites than urban citizens (Katz et al., 2020; Pritchard, 1935; Rader et al., 2020; Scoglio et al., 2010). In addition, rural populations have significantly different incidences of comorbidities including diabetes, obesity, and respiratory conditions that may impact susceptibility during infectious disease outbreaks (Garcia et al., 2017). Ultimately, these factors are associated with significantly different rates of complications from infectious disease outbreaks at the county level over the past 40 years (El Bcheraoui et al., 2018).

Currently, there are limited studies assessing the immunological response to COVID-19, and frequency of SARS-CoV-2 in rural areas. The goal of this research was to assess the frequency and characteristics of SARS-CoV-2 infection in a rural population. This study sampled 150 participants from Montgomery, WV a rural community with a population of 1,592 individuals in 2019. Serological analyses of SARS-CoV-2 exposure were conducted to understand binding antibody levels, neutralizing antibody levels and demographic infection distributions in this rural West Virginia populace. With the knowledge that rural states like West Virginia exhibited different rates of SARS-CoV-2 infection throughout the pandemic, and that these areas have high incidences of comorbidities, we aimed to better understand the serological hallmarks of SARS-CoV-2 exposure in rural citizens.

## METHODS

### Participant enrollment

Study recruitment was conducted on September 10, 2020 at Montgomery General Hospital. After reviewing the informed consent document with any questions answered by onsite study personnel, those desiring to participate in the study signed the informed consent document. Medical records of consenting patients were obtained in addition to self-reported age, biological sex, ethnicity, race, medication list, SARS-CoV-2 test dates (non-specified as PCR or antigen tests) with results, and whether they had experienced any viral-like illness symptoms throughout the time period of the pandemic. Consenting patients were then admitted to a separate room for venipuncture. This study was completed and overseen by the provisions of WVU IRB#: 2008077079.

### Serum sampling of study participants

Approximately 10 ml of blood was obtained from consented patients via venipuncture, performed by licensed medical staff. Blood was collected into Tiger-top blood tubes, and specimens were centrifugated at 3500rpm for 15 minutes (Hettich Rotina-380) and then transported on dry ice to the WVU Vaccine Development center. Serum was aliquoted from the blood collection tubes into barcoded 13mm sample tubes and frozen at -80°C until use.

### Quantitative analysis of anti-SARS-CoV-2 IgG production

Anti-SARS-CoV-2 RBD IgG levels were determined by automated ELISA using the Tecan Freedom EVO 150/200 series. All samples were screened for anti-SARS-CoV-2 activity by diluting 40µL of serum samples into 160µL of 1% non-fat milk dissolved in Phosphate Buffered Saline (PBS) + 0.1% Tween 20 in a non-binding 96-well plate by a Tecan Freedom EVO 150. Diluted specimens were further serially diluted (5-fold) down the columns of the plate by the Tecan Freedom EVO 150. Samples (100µL) were transferred to a 96-well high-binding plate (Pierce Part #:15041) coated with SARS-CoV-2 RBD (2µg/mL produced as previously described(Horspool et al., 2021)) and blocked with 3% milk diluted in PBS +0.1% Tween 20 as described previously(Horspool et al., 2021) by a Tecan Freedom EVO 200. Plates were incubated at room temperature shaking at 60rpm for 10min. After incubating, plates were washed four times with 200uL of PBS + 0.1% Tween 20 by a Tecan HydroSpeed attached to the Tecan Freedom EVO 200. Secondary antibody (100µL of 1:500 goat-anti-human IgG HRP Invitrogen Part #: 31410) was then added to each well. Plates were incubated at room temperature shaking at 60rpm for 10min. After incubation, plates were washed five times with 200uL of PBS + 0.1% Tween 20 by a Tecan HydroSpeed attached to the Tecan EVO 200. After washing, TMB (100µL of 1:1 reagent A : reagent B, Biolegend Part #: 421101) was added to the each well of the plate. Plates were incubated at room temperature for 10min without shaking in the dark. Stop solution (25µL of 3M hydrochloric acid) was added to stop the reaction. Plates were then read by a Sunrise plate-reader and absorbance in each well was measured at a wavelength of 450nm. Data analysis was completed in GraphPad Prism 9. Samples were run simultaneously on the Abbott Architect system as a commercial qualitative comparison of serological results.

### Culture of B. pertussis for ELISA

*B. pertussis* (UT25) was struck on Bordet Gengou agar (Remel™ R45232) supplemented with 15% defibrinated sheep blood (Hemostat Laboratories) and incubated for 2 days at 37°C. After two days, bacteria from plates were swabbed using polyester swabs (Fisher Scientific 22-029-574) into complete Stainer-Scholte Medium (SSM) in new 125mL flasks. Cultures were incubated at 37°C for 24 hours shaking at 180rpm. After incubation, *B. pertussis* cultures were diluted to an OD_600_= 0.24 and used to coat ELISA plates.

### Analysis of anti-B. pertussis and anti-Pertussis toxin IgG levels

High binding ELISA plates (Pierce 15041) were coated with *B. pertussis* (50µL of OD=0.24 *B. pertussis* in Stainer-Scholte Medium) or Pertussis toxin (List Labs #180 at 1µg/mL). Plates were incubated overnight at 4°C. Plates were washed 3 times with 200µL of PBS + 0.1% Tween 20. Blocking buffer (200µL of 5% non-fat milk) was added to each well and plates were incubated for 1hr at 37°C. Plates were washed 3 times with 200µL of PBS + 0.1% Tween 20. Serum samples (25µL) from participants were added into 1% non-fat milk dissolved in PBS + 0.1% Tween 20 in the first row of the plate. Samples were serially diluted (5-fold dilution) down the plate to row G. Row H contained PBS as a negative control. Plates were incubated for 2hrs at 37°C. Plates were washed 4 times with 200µL of PBS + 0.1% Tween 20. Secondary antibody (100µL of 1:500 goat-anti-human IgG HRP Invitrogen Part #: 31410) was then added to each well. Plates were incubated for 1hr at 37°C. After incubation, plates were washed five times with 200uL of PBS +0.1% Tween 20. TMB (100µL) was added to the each well of the plate. Plates were incubated at room temperature for 30min in the dark. Stop solution (25µL of 3M hydrochloric acid) was added to stop the reaction. Plates were then read by a Sunrise plate-reader and absorbance in each well was measured at a wavelength of 450nm.

### Quantification of neutralizing antibodies

An assay to assess neutralizing antibody (nAb) levels was developed using Luminex bead and Magpix technologies. SARS-CoV-2 RBD (1µg) produced at WVU as described previously(Horspool et al., 2021) was conjugated to Luminex MagPlex® Micospheres (MC10012-YY) using the Luminex xMAP antibody coupling kit (Luminex 40-50016) per the manufacturer’s instructions. Conjugated beads (50µL containing 2000 beads suspended in 1x PBS-TBN diluted from 5x PBS-TBN Teknova #:P0211) were loaded into black non-binding Greiner 96-well plates (Greiner 655900). Serum samples 25µL form participants were added into 100µL of PBS in the first row of a second black non-binding plate. Samples were serially diluted (5-fold dilution) down the plate to row G. Row H contained PBS as a negative control. Diluted serum samples (50µL) were added to the 96-well plate containing the beads, creating a total reaction volume of 50µL beads (2000 beads), 50µL diluted serum. The plates were covered with foil and shaken at 700rpm for 1hr at room temperature. After shaking, beads were pelleted on a 96-well plate magnet and washed two times for two minutes with 200µL of 1x PBS-TBN. Beads were pelleted on the magnet and the wash solution removed. ACE-2-biotin (100µL at 0.25µg/mL, Sino Biological Inc #: 10108-H08H-B) was added to each well. Plates were covered with foil and shaken at 700rpm for 1hr at room temperature. After shaking, beads were pelleted on a 96-well plate magnet and washed two times for two minutes with 200µL of 1x PBS-TBN. Beads were pelleted on the magnet and the wash solution removed. Streptavidin-phycoerythrin (MOSS INC: SAPE-001) (100µL at 4µg/mL) was added to each well. Plates were covered with foil and shaken at 700rpm for 30min at room temperature. After shaking, beads were pelleted on a 96-well plate magnet and washed two times for two minutes with 200µL of 1x PBS-TBN. Beads were resuspended in 100µL of 1x PBS-TBN and analyzed on a Luminex Magpix. Median fluorescent intensity values were plotted against serum dilution factor, and a sigmoidal regression line was fitted to the data using GraphPad Prism. Calculated IC50 values of the sigmoidal curves were plotted separately as a measure of neutralizing capacity.

### Statistical analyses

Statistical analyses were conducted using GraphPad Prism (version 9.0.0). Analyses of data linearity were completed by using the linear regression function in GraphPad Prism. Correlations within datasets were investigated using two-tailed Pearson correlation analyses. To assess statistical differences between two groups, a two-tailed Student *t*-tests was used, or a one-way ANOVA followed by Tukey’s multiple comparison test to assess differences between multiple groups. Statistical significance was determined to be *P* < 0.05.

## RESULTS

### Montgomery, West Virginia anti-SARS-CoV-2 RBD IgG levels

One hundred fifty individuals consented to participate in the study, and venipuncture was performed to assess prior exposure to SARS-CoV-2. The mean participant age was 48 years (S.D. = +/- 15 years) and was comprised of 30% biological males (Table 1). SARS-CoV-2 incidence was determined to be 10% (15/150 participants) (Table 1) by combining information from serological testing, patient reporting, and prior PCR based SARS-CoV-2 test results obtained from medical records. Quantitative serological testing for anti-SARS-CoV-2 RBD IgG was completed at the WVU Vaccine Development center utilizing methodology and cutoff values previously described(Horspool et al., 2021). The WVU VDC antibody analyses identified 87% (13/15) of individuals with medical history of SARS-CoV-2 exposure within six months of this study (Table 1). A similar commercial qualitative SARS-CoV-2 IgG testing method (Abbott) identified 67% (10/15) exposed individuals.

**Table 1.** Study participant information: Average age, biological sex distribution, and SARS-CoV-2 exposure were determined for the participants of this study. Proportion of patients exposed or not exposed to SARS-CoV-2 and the WVU VDC detection rates of known exposed individuals. n = 150 participants.

| Demographics | Age | Proportion biologically male | SARS-CoV-2 +/- |
| --- | --- | --- | --- |
| Average | 48 yrs | 30% | 9% |
| SD | 15 yrs | N/A | N/A |
| SARS-CoV-2 history | Positive | Negative | VDC Detected |
| # of total | 15/150 | 135/150 | 13/150 |
| % of total | 10 | 90 | 8.67 |

### Anti-SARS-CoV-2 IgG levels decrease post-exposure

Serum from participants was diluted on direct-ELISA plates to determine semi-quantitative values of anti-SARS-CoV-2 RBD IgG production. Absorbances of diluted patient samples and a comparison of area under the curve analysis described previously(Horspool et al., 2021) demonstrate clear differences between SARS-CoV-2 exposed and non-exposed participants (Figure 1A-B). Among those previously infected with SARS-CoV-2, neither age nor sex were associated with timing of infection (Supplementary Figure 1). To understand the impact of time on anti-SARS-CoV-2 RBD IgG levels, anti-RBD AUC values of exposed individuals were correlated against time post-SARS-CoV-2 exposure (Figure 1C). There is a significant inverse correlation between quantity of anti-SARS-CoV-2 RBD IgG and days post-infection, and individuals who were admitted to the hospital during infection had higher anti-RBD IgG levels. Data of anti-SARS-CoV-2 IgG levels over time were compared to a previous study of acute SARS-CoV-2 IgG levels in a West Virginian hospital(Horspool et al., 2021). Irrespective of hospitalization, individuals infected with SARS-CoV-2 within 3 months of diagnosis produced similar quantities of anti-SARS-CoV-2 RBD levels (Figure 1D). These levels significantly decreased over subsequent months post-exposure.

**Figure 1.**
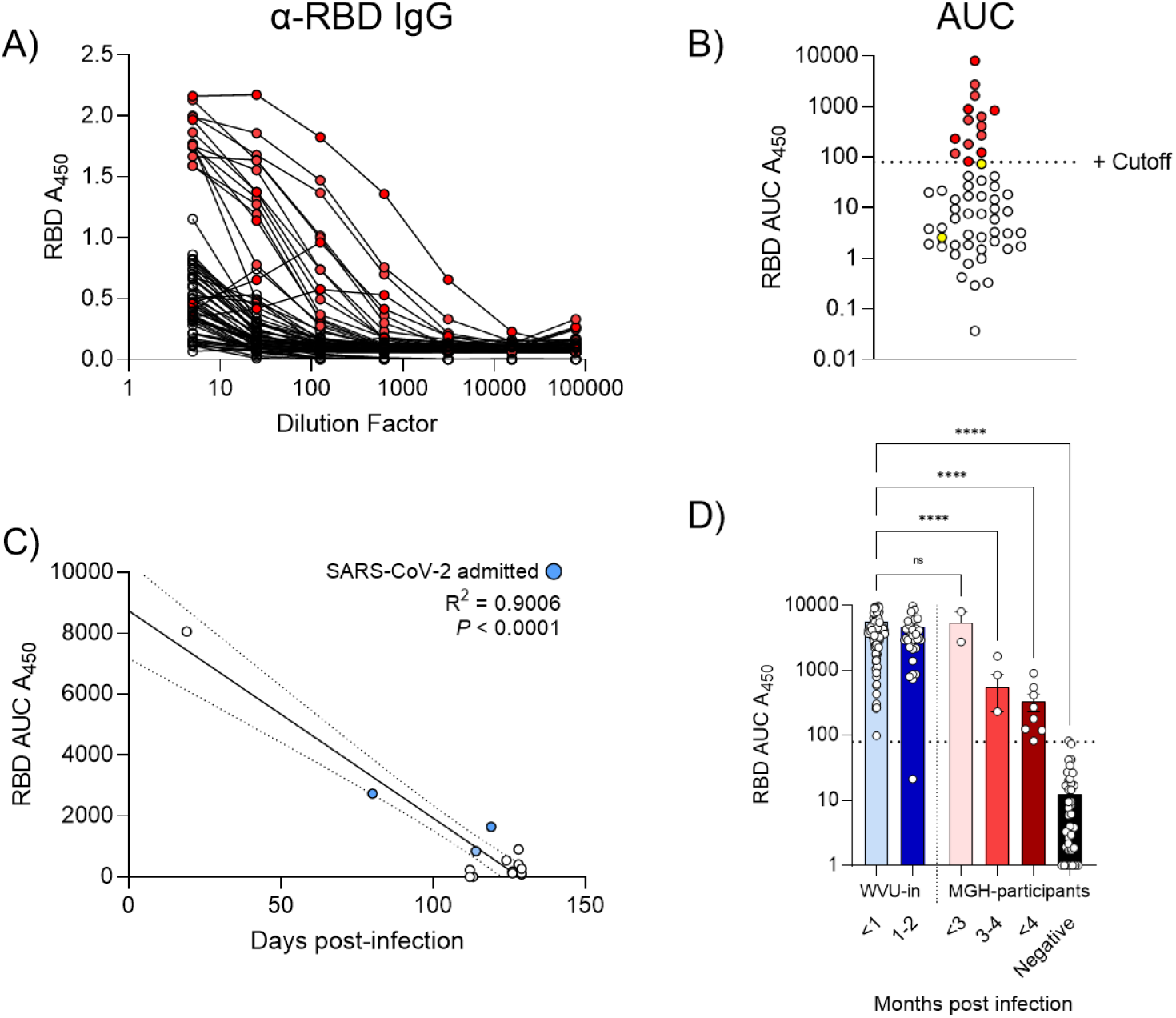
Anti-SARS-CoV-2 IgG levels of study participants: Levels of anti-SARS-CoV-2 IgG were determined for study participants. A) Titration of patient samples on SARS-CoV-2 ELISA plates. Red symbols = SARS-CoV-2 exposed, white symbols = non-exposed, yellow symbols = not detected by WVUVDC). B) Area under the curve (AUC) analysis of each patient curve plotted against previously defined cutoff values for SARS-CoV-2 exposure^13^. C) Anti-SARS-CoV-2 RBD IgG levels of exposed participants were plotted against the number of days after the participant tested positive for SARS-CoV-2. Blue dots represent participants that were admitted to the hospital during infection. D) Comparison of anti-RBD AUC values from a prior study and this study over time. Statistical significance was assessed by a two-tailed Pearson correlation or a one-way ANOVA followed by a Tukey’s multiple comparison test. **** = *P* < 0.0001, ns = not significant.

### Study participants possess normal immunogenic profiles against B. pertussis

Immunocompromised individuals can lack appropriate responses to infectious diseases and vaccines. These individuals may bias results in a serological study. To understand whether the SARS-CoV-2 exposed participants from this study possessed abnormal immunogenic profiles, we assessed antibody production to an additional pathogen most people are vaccinated against: *B. pertussis*. Serum from SARS-CoV-2 exposed and a selection of non-exposed individuals were tested for IgG production against whole *B. pertussis* and Pertussis toxin. The data indicate that there was no difference in anti-*B. pertussis* (Figure 2A-B) or anti-Pertussis toxin (Figure 2C-D) IgG levels between individuals exposed or not exposed to SARS-CoV-2. These data suggest that there is a similar immunogenic profile between each cohort and can assume this study was mostly comprised of immunocompetent subjects.

**Figure 2.**
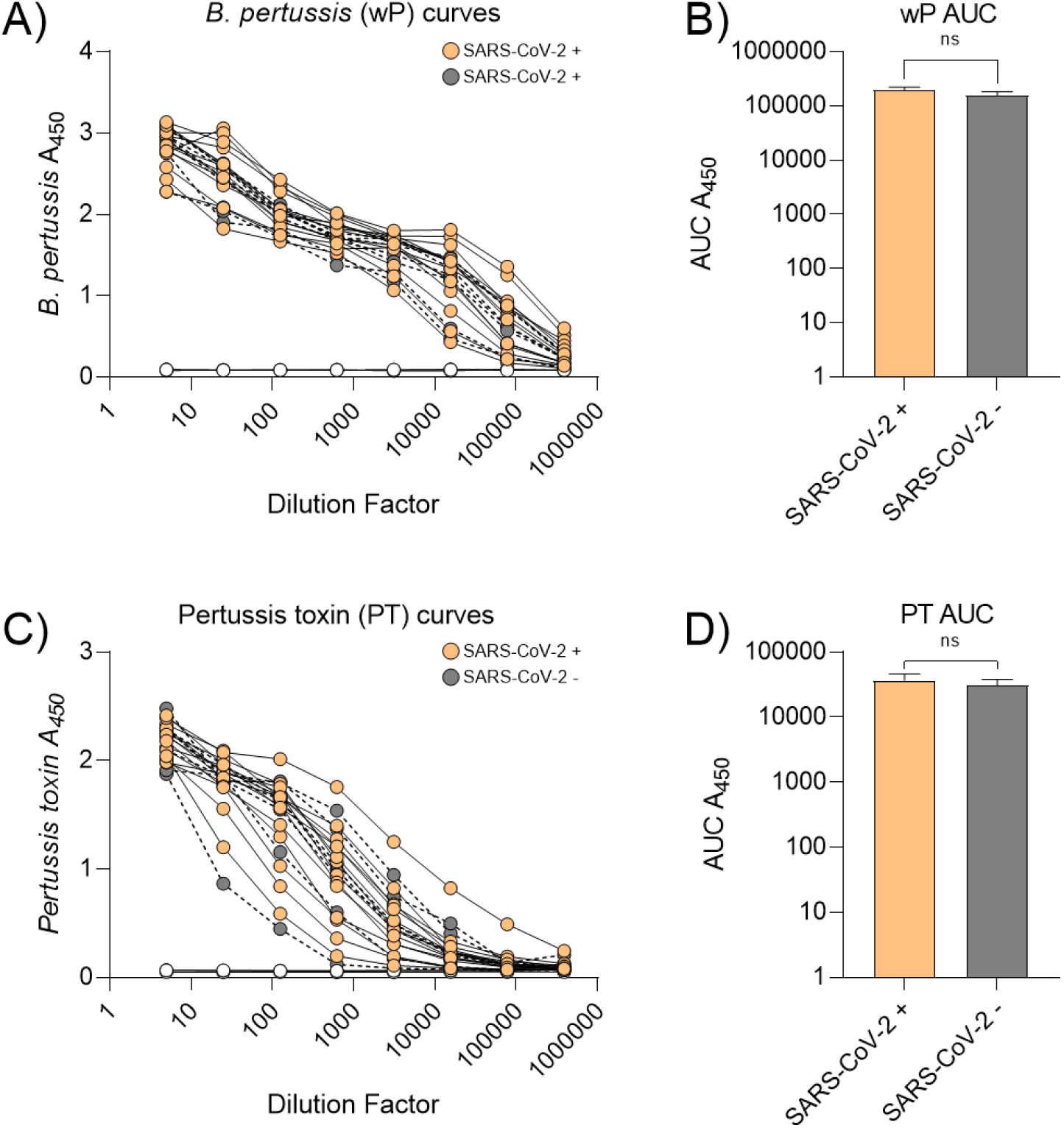
Anti-*B. pertussis* and anti-Pertussis toxin IgG levels of study participants: A) Titration of participant samples on *B. pertussis* ELISA plates. Orange symbols = SARS-CoV-2 exposed, grey symbols = non-exposed, white symbols = negative control). B) Area under the curve (AUC) analysis of *B. pertussis* ELISA curves of participants exposed or not exposed to SARS-CoV-2. C) Titration of participant samples on Pertussis toxin ELISA plates. Orange symbols = SARS-CoV-2 exposed, grey symbols = non-exposed, white symbols = negative control). D) Area under the curve (AUC) analysis of Pertussis toxin ELISA curves of participants exposed or not exposed to SARS-CoV-2. Statistical significance was assessed by a two-tailed student’s t-test. ns = not significant.

### Neutralizing antibodies against SARS-CoV-2 decrease post-exposure

Antibodies that neutralize the capacity of SARS-CoV-2 RBD to bind to the human ACE-2 receptor were evaluated from participant serum. Neutralizing antibodies (nAbs) blocking an interaction between immobilized RBD and soluble ACE-2 with a fluorescently labeled epitope were quantified by a modified Luminex assay. Neutralization curves and IC50 values of participant serum indicate that approximately 53% (8/15) of SARS-CoV-2 exposed individuals have nAbs that are sufficient to block the RBD-ACE-2 interaction *in vitro*. nAb quantities were further investigated by correlating nAb IC50s to time post-SARS-CoV-2 exposure. IC50 values appear to decrease significantly over five months post-exposure (Figure 3C) particularly in individuals greater than 3 months post-exposure (Figure 3D).

**Figure 3.**
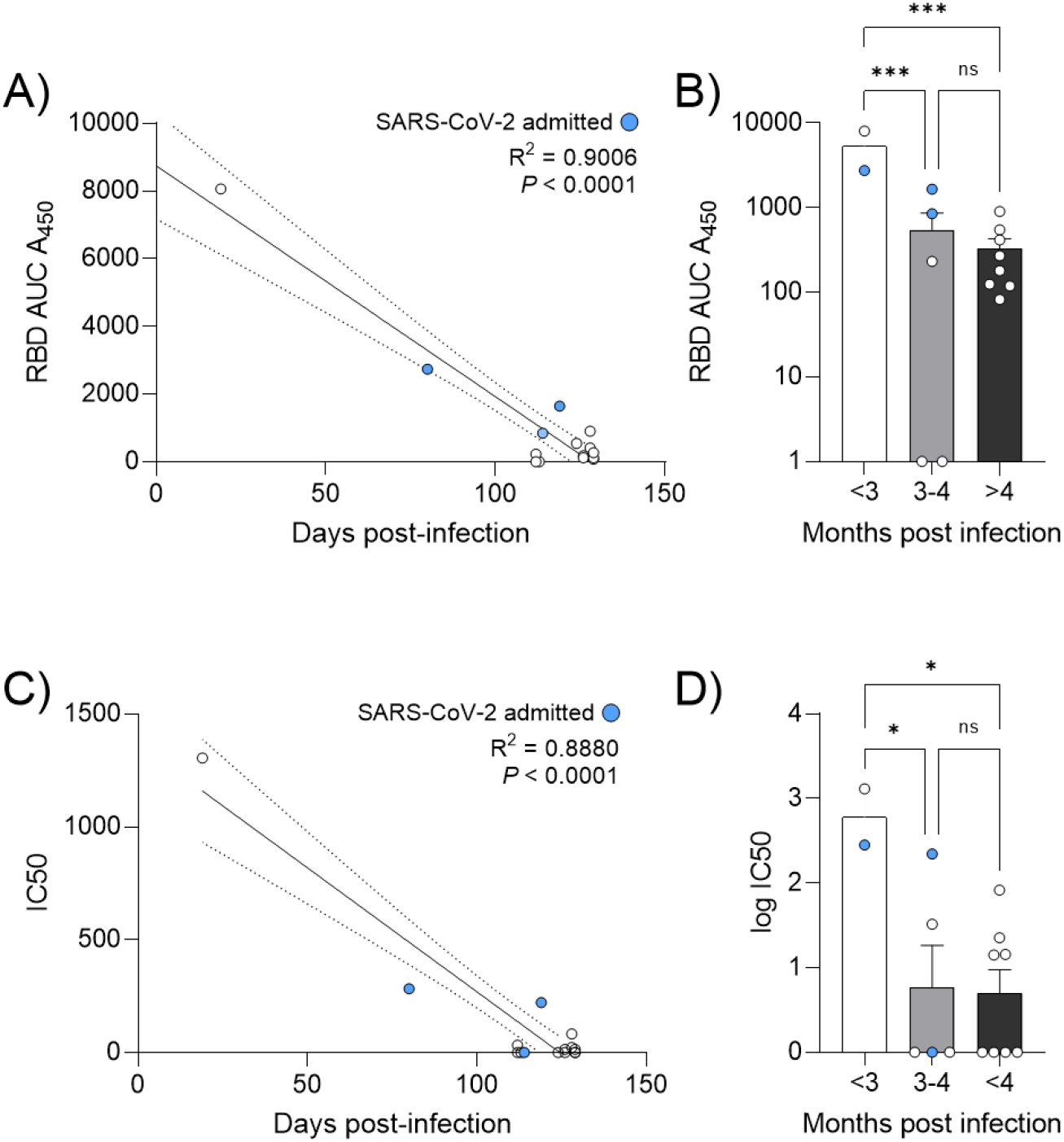
Anti-SARS-CoV-2 IgG levels appear decreased over time: A) Anti-SARS-CoV-2 nAb curves of study participants. B) Anti-SARS-CoV-2 nAb IC50 values at the time of this study were plotted and a cutoff between neutralizing and non-neutralizing samples was drawn. C) nAb levels of exposed participants were plotted against the number of days after the participant tested positive for SARS-CoV-2. Blue dots represent participants that were admitted to the hospital during infection. D) Comparison of nAb IC50 values from this study over time. Linear regression was calculated in GraphPad Prism 9. Statistical significance was assessed by a two-tailed Pearson correlation or a one-way ANOVA followed by a Tukey’s multiple comparison test. * = *P* < 0.05, ns = not significant.

### Correlative analyses of demographics and symptom reporting

Anti-SARS-CoV-2 RBD IgG levels of SARS-CoV-2 of non-hospitalized individuals greater than 3 months post-exposure were plotted against participant age and biological sex. There was no significant correlation between participant age (Figure 4A) or participant biological sex (Figure 4B) and anti-SARS-CoV-2 RBD IgG levels. Individuals less than 3 months post-exposure or that were hospitalized had significantly higher levels of IgG and introduced bias un-related to the demographic being evaluated. However, there was a noticeable difference in reporting of viral-like symptoms between exposed individuals when compartmentalized by age (Figures 4C-E). Participants under 30 years old (Figure 4C) reported viral-like symptoms at a lower frequency than individuals greater than 30 years old (Figures 4D-E).

**Figure 4.**
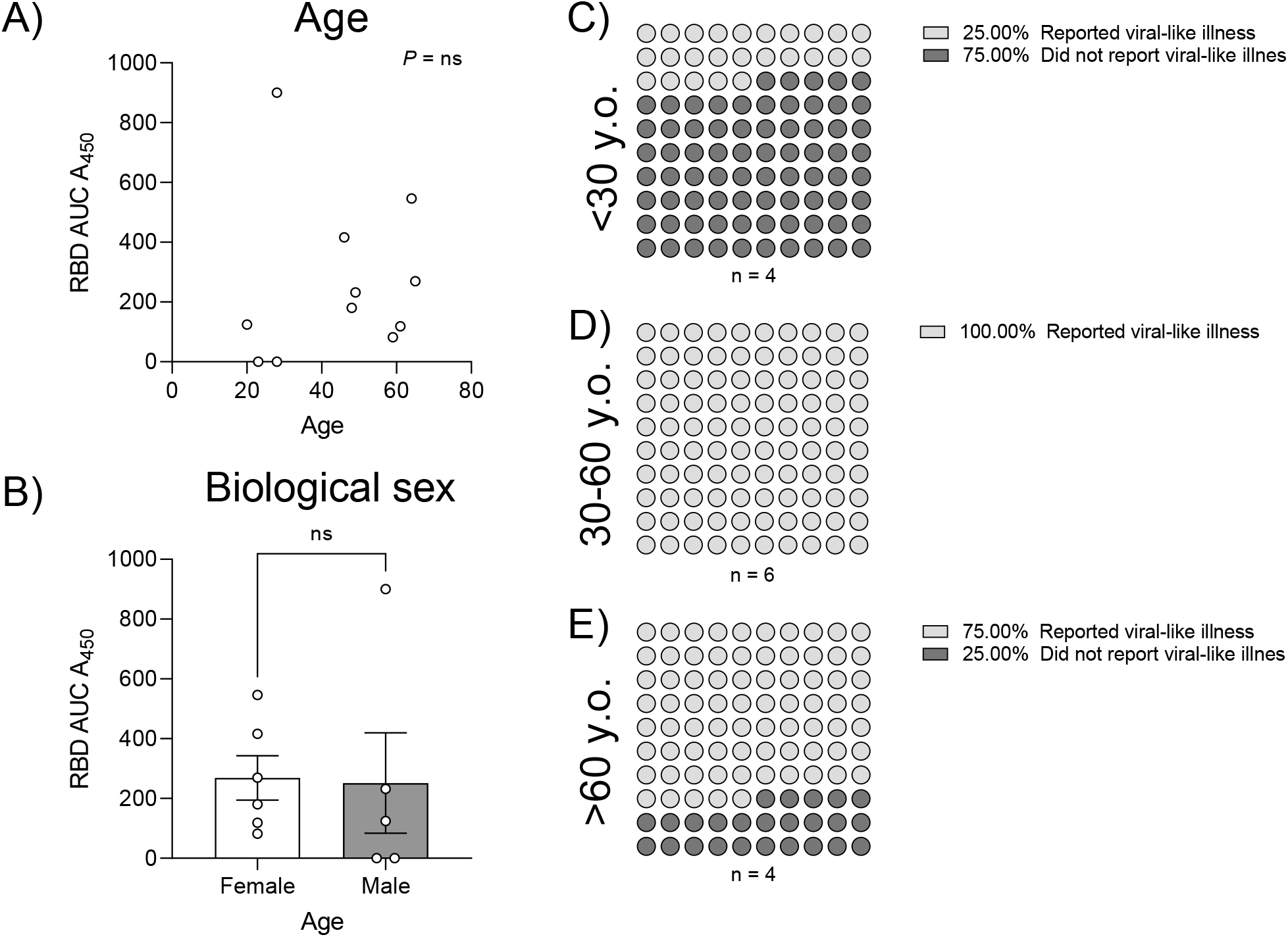
Correlations between anti-SARS-CoV-2 IgG serological data and questionnaire responses: A) Age and B) biological sex were plotted against anti-SARS-CoV-2 RBD IgG levels. Frequency of reporting viral-like illness symptoms from SARS-CoV-2 exposed participants C) <30 years old, D) 30-60 years old, and E) >60 years old. Statistical analyses were completed by Pearson correlation (age) or two-tailed Student’s t-test (biological sex). ns = not significant.

## DISCUSSION

Epidemics and pandemics impact rural and urban communities differently. Community health, disease transmission dynamics, and resources vary in large cities vs rural towns(Mueller et al., 2021; Schweda et al., 2021; Scoglio et al., 2010; Singh et al., 2019). This study revealed that the incidence of SARS-CoV-2 in a subset of a rural West Virginian population is approximately 10% which is high relative to the national average for that time period(Johns Hopkins University of Medicine, 2021). It is possible that the decreased number of community facilities including convenience stores, food services, hospitals, etc. may have increased the proportion of the community contacted by an infected person. In addition, rural communities are statistically more likely to have residents with comorbidities including diabetes, obesity, and respiratory conditions(Garcia et al., 2017). Increased incidence of comorbidities may impact the likelihood of contracting COVID-19 in a rural setting, though larger controlled studies must be completed to address this hypothesis.

The significant decreases observed in anti-SARS-CoV-2 IgG over time have been described by others previously(Marot et al., 2021; Perreault et al., 2020; Self et al., 2020; Ward et al., 2020), but not in the context of rural communities. Declining anti-SARS-CoV-2 antibody levels is a cause for concern for individuals that have convalesced from infection and warrants further investigation. The comparable trend of declining nAbs in participants is particularly alarming as this indicates that this decline may not be limited to IgGs but may encompass other classes of antibodies interfacing with SARS-CoV-2 RBD. Additional results have been produced by other studies internationally with variable rates of decline (Choe et al., 2021; Long et al., 2020; Seow et al., 2020; Wang et al., 2020). Ultimately, these data may be critical in understanding the risks of declining anti-SARS-CoV-2 immunity and the potential for SARS-CoV-2 reinfection in rural communities. Further studies examining this effect in other areas in the United States will be imperative to better understand differences in urban-rural immunity as the pandemic continues.

Age and sex have been determined to have significant effects on some anti-SARS-CoV-2 IgG production (Chakravarty et al., 2020; Horspool et al., 2021; Robbiani et al., 2020; Scully et al., 2020; Takahashi et al., 2020; Zeng et al., 2020). In many of these studies, neither age, nor sex significantly impact anti-SARS-CoV-2 RBD IgG production, a result that is reflected in this study. However, the data presented here is limited in the number of positive cases identified due to a sampling of a small rural community. Despite this finding, an interesting trend emerged in reports of viral-like symptoms from individuals exposed to SARS-CoV-2. The incidence of reported viral-like illness symptoms in participants over the age of 30 years old indirectly suggests a more severe course of COVID-19. It has been established that COVID-19 severity is increased with age (Cortis, 2020; Davies et al., 2020; Yuki et al., 2020). Indeed, three individuals in this study were hospitalized for COVID-19 all of whom were over the age of 30 and two of whom were over the age of 60. Although much larger studies have assessed these variables in urban populations, it is informative to note that similar conclusions may be applicable to this rural population.

Overall, this study provides a compact analysis of SARS-CoV-2 incidence conducted at a rural West Virginia hospital. Total incidence taken at this community hospital appears high relative to the national average at that time which may be due to different rural transmission networks. The levels of anti-SARS-CoV-2 RBD IgG and nAbs in participants appear to decrease over six months post-infection, and no participants appeared to have abnormal immunogenic profiles to a standard vaccine pathogen. The trends described in this work are independent of age and sex in concordance with other literature. Overall, the results of this study suggest decreasing immunity in rural persons which may negatively impact ability of a population to attain herd immunity without a vaccine that provides longer-lasting protection. This emphasizes the need for vaccine coverage among a large portion of the population. Ultimately, this study aids in understanding the manifestation of SARS-CoV-2 in rural populations which may be critical for combatting transmission in underserved communities and may better inform prevention and vaccination efforts.

## Data Availability

Data are available upon request should be noted somewhere in the materials and methods so just add that

## ACKNOWLEDGEMENTS

This project was supported by the Vaccine Development Center at the West Virginia University Health Sciences Center. F.H.D. and the VDC are supported by the Research Challenge Grant no. HEPC.dsr.18.6 from the Division of Science and Research, WV Higher Education Policy Commission. Research reported in this publication was also supported by the National Institute of General Medical Sciences of the National Institutes of Health under Award Number 5U54GM104942-05. The content is solely the responsibility of the authors and does not necessarily represent the official views of the National Institutes of Health.

## AUTHOR CONTRIBUTIONS

AMH, RE, AS, LC, SW, TK, SH, FHD designed the study and planned execution. RE, AS, LC, SW planned resource allocation and venipuncture draws at Montgomery General hospital. MAW, JK, CBB, JH, TYW, MAD, GB, JRB consented patients. AMH processed serum samples. AMH and BPR ran serological tests including ELISAs and neutralization assays. BPR assisted in running ELISAs. All authors contributed to writing and revision of the manuscript.

---

**Supplementary Figure 1.**
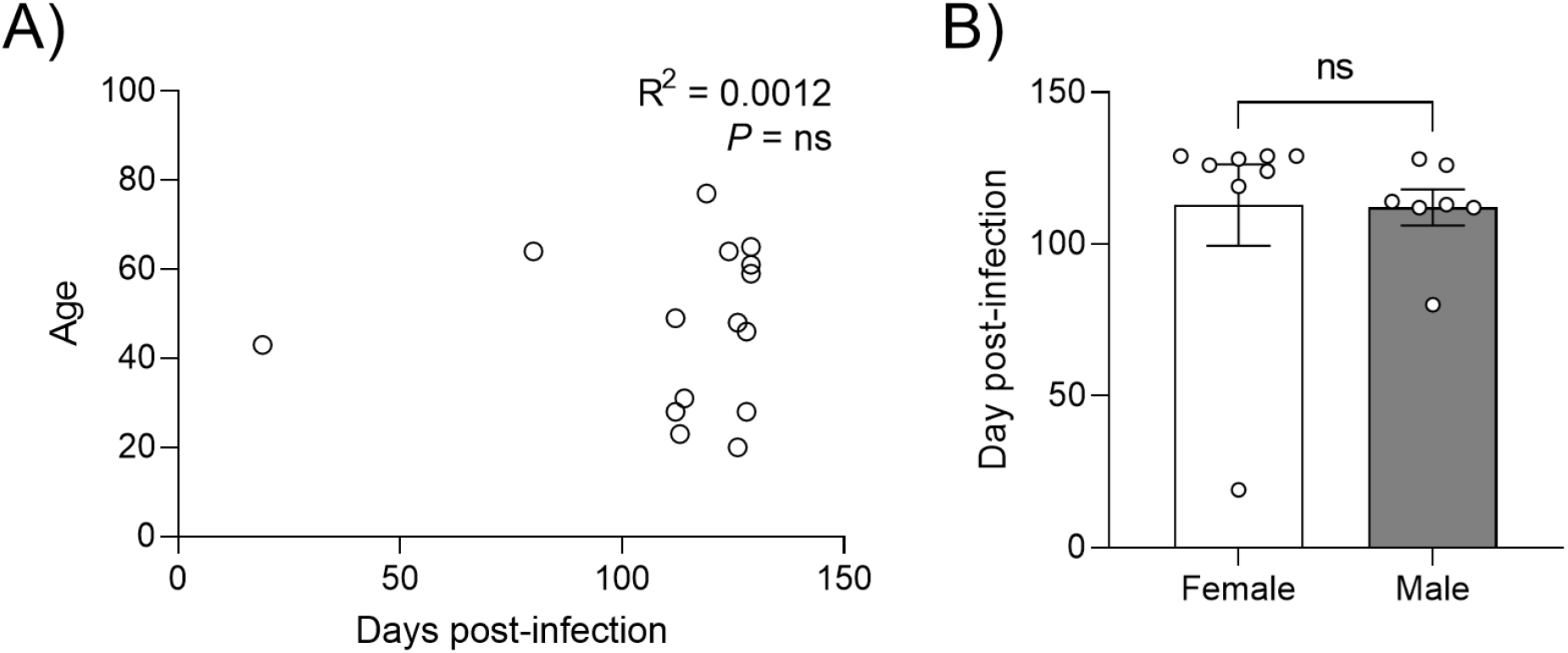
No correlations between age or biological sex and days post-infection: A) Age correlated to days post-infection with SARS-CoV-2 and B) biological sex correlated to days post-infection. Statistical significance was assessed by either two-tailed Pearson correlation or two-tailed student t-test. ns = not significant.

